# Antecedents and outcomes of a late attention deficit hyperactivity disorder (ADHD) diagnosis in females

**DOI:** 10.1101/2025.07.07.25330613

**Authors:** Joanna Martin, Olivier Y. Rouquette, Kate Langley, Miriam Cooper, Kapil Sayal, Tamsin Ford, Ann John, Anita Thapar

## Abstract

Females receive an attention deficit hyperactivity disorder (ADHD) diagnosis at an older age than males. We examined the antecedents and outcomes of later (age 12+) diagnosis in females using data from a Welsh nation-wide electronic cohort of 13,593 individuals (N=2,680 (19.7%) females) with ADHD and 578,793 individuals (N=286,734 (49.5%) females) without ADHD. We compared females with later diagnoses (ages 12–25) to those with earlier, timely diagnoses (ages 5–11) and no diagnosis, in terms of childhood (ages 5–11) antecedents and adolescent/adult (ages 12–25) outcomes. We also tested for sex differences. Females with later diagnosed ADHD used more adolescent/adult healthcare services and had worse mental health, educational and socioeconomic outcomes than females diagnosed earlier. Health and educational difficulties were already evident in childhood in this group. Many outcomes were exacerbated in females compared to males. Timely childhood ADHD diagnosis is necessary to mitigate later risks, especially for females.

## Introduction

Attention deficit hyperactivity disorder (ADHD) is a common neurodevelopmental condition that is less likely to be diagnosed in girls/women (hereon: females) compared to boys/men (hereon: males)^1^, partly due to under-recognition^2^. When ADHD is recognised in females, it is diagnosed on average at a later age than in males^3–5^. It is well-evidenced that ADHD is associated with mental health conditions and poorer social, educational and occupational functioning. A delay in diagnosis, meaning that individuals miss out on timely support and treatment for their ADHD, is likely to exacerbate these adverse outcomes. A better understanding is needed of factors that influence ADHD diagnosis timing and the potential health and wellbeing consequences of delayed diagnosis, especially in females. This is a necessary step towards earlier recognition of ADHD in this historically overlooked group.

It is well established that being female is an important barrier to accessing healthcare related to ADHD^6^. Other barriers to ADHD diagnosis and accessing treatment and support include low socioeconomic status, ethnic minority status, and the intersection of these identities (Madsen et al. 2018; Mennies et al. 2020; Wright et al. 2015). Females with ADHD are at higher risk for a variety of co-occurring neurodevelopmental and psychiatric conditions, receive more psychotropic medication prescriptions, and show higher healthcare use, compared to males with ADHD^3,7–10^. Further understanding of potential barriers to accessing services, the antecedents of late ADHD diagnosis, and how these vary by sex, are essential to improve timely assessment of ADHD and subsequent clinical care for underserved populations and thereby to reduce inequalities. Understanding these could identify early signs which may indicate the need for referral for an ADHD assessment.

The annual UK costs of health, education and social care related to ADHD in adolescents were estimated at £670 million over a decade ago^11^. This figure is likely higher today and does not capture costs to healthcare services and the wider economy associated with potential consequences of delays in diagnosis and treatment. As such, the economic cost of undiagnosed and untreated ADHD is likely to be much higher, particularly in adulthood^12^.

Diagnosed ADHD is associated with substantially increased economic costs related to psychiatric and somatic disorders, drug abuse, injuries, criminal justice, and educational impacts^13–15^. Undiagnosed ADHD further impacts across a range of life domains, including high rates of substance misuse, psychiatric conditions (e.g. depression), criminality, and lower educational attainment^16^. However most studies of undiagnosed ADHD have only involved adults and many were primarily or exclusively about males, with only two studies in children and no studies focusing on females^16^. The childhood studies found associations between undiagnosed ADHD and lower self-esteem, more mental health difficulties (e.g. depression, self-harm), peer relationship difficulties, and hospital admissions for injuries, with no investigation of sex differences^17,18^.

Timely recognition is important because ADHD treatment improves long-term social, educational and occupational outcomes, and reduces the risks of depression, suicide, substance abuse, accidents, and criminality^19–21^. Given the later average age at ADHD diagnosis in females^3–5^, we need to consider whether the consequences of later diagnosis and treatment on wellbeing, health, and educational outcomes are exacerbated in females compared to males.

The objective of this study is to address these knowledge gaps, using a whole-nation electronic register-based cohort of young people in Wales. The specific aims are to examine the association between timing of ADHD diagnosis and: 1) healthcare, educational, and demographic childhood antecedents, and 2) healthcare, educational, and socioeconomic outcomes in adolescence and early adulthood. We compared females with a first recorded ADHD diagnosis in adolescence/adulthood, with females: a) first diagnosed in childhood (timely diagnosis) and b) without ADHD, as well as testing whether results differed in males. We hypothesised that predictors of later diagnosed ADHD in females would include greater healthcare access and educational difficulties in childhood, compared to females without ADHD, but not necessarily to the same extent as females with earlier diagnosed ADHD. We also hypothesised that females with later diagnosed ADHD would exhibit worse healthcare, educational, and socioeconomic outcomes in adolescence and adulthood compared to both females with earlier diagnosis and no ADHD. Sex comparisons were largely exploratory, with the expectation that some outcomes would be exacerbated in females.

## Method

### Data

We used nationwide register data from the Secure Anonymised Information Linkage (SAIL) databank (http://www.saildatabank.com)^22–24^. SAIL is a person-level linkable data repository that covers routinely collected primary and secondary healthcare, education, and social care data for Wales (including approximately 5.45 million individuals). Data were linked using a secure research infrastructure platform, supported by the Adolescent Mental Health Data Platform (https://adolescentmentalhealth.uk), that pools data from multiple sources to create a research cohort using person-specific anonymous linkage field identity numbers, using all records linked deterministically or probabilistically with a linkage score >0.9^25^. Ethical approval for this project was granted by the Information Governance Review Panel (IGRP #1335); this is an independent body consisting of government, regulatory and professional agencies^26^.

The following datasets, covering the whole population of Wales, unless otherwise specified, were linked at an individual level. Information on date of birth and sex assigned at birth was obtained from the Welsh Demographic Service Dataset (WDSD), which is a demographics register of people registered with a general practice in Wales. Data on death was obtained from the Annual District Death Extract (ADDE). The databank includes healthcare records from the Welsh Longitudinal General Practice Database (WLGPD), including attendance and clinical information for primary care data (covering 86% of the population of Wales), the Outpatient Database for Wales (OPDW) including hospital outpatients appointments, the Patient Episode Database for Wales (PEDW), including hospital admissions (inpatients and day cases, including diagnoses), and Emergency Department Dataset (EDDS), including accident and emergency department attendance. These datasets include dates of diagnoses, appointments and prescription information.

Linkage with the identity number of the birth mother was available via the National Community Child Health Database (NCCH), which was used to obtain maternal records of ADHD and depression, covering 81.5% of the study population. Information on ethnicity came from the Office for National Statistics (ONS) Census 2011: Welsh Records (CENW), covering 80.8% of the study population. Information on young people who have had contact with social services came from linked databases held by Welsh Government, including the Children Receiving Care and Support (CRCS), Looked After Children Wales (LACW), and Children in Need Wales (CINW) census data^27^. Information on education (exam results and absences) was obtained from the Pre16 Education attainment in Wales (EDUW) dataset, covering 75.4% of the study population.

### Cohort definition

The study follow-up period used data from 01.01.2000 until 31.12.2019. Individuals were included if they were born between 01.01.1989 and 31.12.2006. Each individual was followed up from their 5^th^ birthday, the start of the study period, start of registration with a GP surgery providing data to SAIL, or start of registration at a Welsh address in the WDSD (whichever came last). The end of each individual’s follow-up period was defined as their 25^th^ birthday, the end of the study period, end of registration with a GP surgery providing data to SAIL, end of registration at a Welsh address, or their date of death (whichever came first). Individuals with less than 365 (non-continuous) days of follow-up data between the ages of 5–18 were excluded.

Sub-samples of this overall cohort were used to ensure sufficient follow up time of individuals in each analysis. For aim 1, which focuses on childhood antecedents, we restricted analyses to individuals followed up for a minimum of 365 days between ages 5– 11. For aim 2, which focuses on outcomes during adolescence and emerging adulthood, we restricted analyses to individuals followed up for a minimum of 365 days between ages 12– 18.

The ADHD sample was defined as those meeting the above inclusion criteria, who have received an ADHD diagnosis or a prescription for any ADHD medication at any time during each individual’s follow up period. ADHD was defined using a list of International Classification of Diseases, 10th Revision (ICD-10) codes relevant to ADHD, that are used in secondary care, as well as Read Code lists and algorithms (version 2), which are a coded thesaurus of clinical terms (including diagnoses, medications, and administrative codes) used in primary care in the UK National Health Service (NHS). These codes have previously been validated using an external cohort of young people with ADHD^28^, with additional codes for ADHD medications included based on consultation with a clinical psychiatrist specialising in ADHD in Wales^3^.

The comparison (non-ADHD) sample included all individuals born in the same time period, with a sufficient amount of follow up data (as defined above), who have never had a recorded ADHD diagnosis or prescription.

### Variable definitions

#### Demographic factors

We examined sex assigned at birth (from WDSD, ethnicity group (ethnic majority or any ethnic minority; sample sizes were too small to define more specific ethnic groups), and socioeconomic deprivation, using the Welsh Index of Multiple Deprivation (WIMD version 2014, based on average income, education, unemployment, and health statistics in a local geographical area) at the start of entry into the study. Quintiles of deprivation were defined using the national WIMD cut-offs (1=least deprived, 5=most deprived). We also examined socioeconomic deprivation index at the end of each individual’s follow up period, in those who were aged 18 or older by the end of the study. We defined a subgroup of individuals who have ever experienced social services involvement, including those who have been looked after in state/foster care, based on the LACW dataset, as well as whether they have been listed on the child protection register, based on a combination of the CRCS and CINW datasets^27^.

#### ADHD diagnosis and medication

Age at first recorded ADHD was defined using the first date of ADHD diagnosis or prescription recorded in either primary or secondary care records (WLGPD/PEDW). Because the precise age of first recorded ADHD could be influenced by numerous factors, including waiting list variability across geographical location, we generated binary variables related to diagnosis timing. Earlier ADHD diagnoses were defined as those that were first recorded before age 12 years (i.e. prior to or around the time of the key life transition from primary to secondary school in Wales and in line with the ADHD symptom age at onset definition used by the DSM-5). Later diagnoses were defined as those first recorded on or after an individual’s 12^th^ birthday (i.e. during secondary school or emerging/young adulthood).

#### Other conditions and medications

Lists of previously published Read Codes validated through the Adolescent Mental Health Data Platform were used to define mental health conditions and medication prescriptions, for the following conditions: autism, specific learning difficulties, and conduct disorder^28–30^, anxiety and depression^31^, self-harm (including non-fatal intentional self-harm, self-injury, self-poisoning and suicide attempts)^32,33^, alcohol and psychoactive (excluding tobacco) substance use (including harmful use, dependence, or use resulting in hospital admission)^32^, eating disorders (including anorexia nervosa, bulimia nervosa, and ‘eating disorders, not otherwise specified’)^34^, bipolar disorder and schizophrenia/other psychotic disorder^35^. We also used previous algorithms of Read Codes to derive variables for the following prescription categories: anti-anxiety (including anxiolytics, sedatives, and hypnotics)^31^, antidepressant^31^, and antipsychotic^36^ medication prescriptions. Separate binary variables were defined for whether a specific condition/prescription was first recorded prior to age 12 (aim 1) or first recorded at age 12+ (aim 2).

#### Healthcare service use

Variables were defined for the numbers of recorded primary care, outpatient, inpatient, and emergency department visits per year that each individual was included in the study period (i.e. mean visits per year), split by age (5–11 or 12–18). Due to long tails, these variables were capped at 5 SD away from the mean.

#### Pregnancy

For females, we defined a binary variable for teenage (i.e. age 12–18 years) pregnancy based on previous research^37^, not including early terminated/miscarried pregnancies due to lack of available data.

#### Education

Linked data from the Welsh Government EDUW dataset were used to derive variables related to educational attainment (at key stages 1–4) and absenteeism. Binary variables were derived for whether each individual who undertook the key stage exams passed (coded 0) or failed (coded 1), using data from each person’s last attempted assessment. Key stage (KS) assessments in core subjects are completed in Wales at ages 7/8 (KS1), 11/12 (KS2), 14/15 (KS3), and 16/17 (KS4). KS4 passes were then dichotomised into low or high (5 A*–C) attainment. Records for school attendance were available for academic years starting 2007–2008 to 2018–2019 (N.B. the grading system in Wales did not change the way it did in England during this time period). Persistent absences were defined as a binary variable, per pupil, per year, coded as yes if >10% of possible sessions were missed and no if fewer sessions were missed, in line with previous research, based on current educational guidance^38^. Aggregate binary variables were calculated for the occurrence (yes/no) of any persistent school absences in years when the child was aged <12 years old at the start of each school year and when they were 12 years or older.

#### Maternal diagnoses

For each individual, we also defined variables for any ADHD and depression in the birth mother. Information on these conditions was based on records in primary and secondary care, as defined above. ADHD was considered present if any records were identified at any time point and depression was considered present only if it was during the child’s available follow-up period when they were 5–11 years old. Data from mothers were only included if the maternal follow-up period was a minimum of 365 days during the study period.

### Data analyses

Age at first recorded diagnosis of ADHD was used to dichotomise the sample into those diagnosed earlier (i.e. childhood, between 5–11 years old) and those diagnosed later (i.e. adolescence/early adulthood, between 12–25 years old).

#### Childhood antecedents

We first examined factors during childhood (prior to age 12) that may precede or coincide with the ADHD diagnostic process in early childhood, including specific neurodevelopmental and mental health conditions and prescriptions that are common in childhood, healthcare service use, primary school assessments (KS1 & KS2), school absences, and maternal ADHD or depression. We also examined demographic factors which may be associated with the diagnostic process, including ethnicity, social services involvement, and socioeconomic deprivation.

#### Adolescent/adult outcomes

To determine the impact of timing of ADHD diagnosis, we focused on outcomes during adolescence (ages 12–18) and emerging adulthood (ages 18–25). We examined mental health conditions and prescriptions first recorded at age 12+, healthcare service use, teenage pregnancy (12–18, females only), secondary school assessments (KS3 & KS4), school absences, and socioeconomic deprivation index at study end (in those 18+).

#### Analyses

For both aims, we compared females with a later ADHD diagnosis with females: a) with an earlier diagnosis and b) without ADHD. To determine whether any associations were stronger in females, we analysed males separately and also analysed females and males together, to test for moderation by sex using an interaction test.

All analyses were performed in R-4.4.1 using logistic or linear regression, co-varying for birth year and total study follow up time, to account for variation in dates of birth and length of individual time in the study. Cluster robust standard errors were estimated to account for known siblings, using maternal ID as the clustering variable. All analyses were corrected for multiple testing using false discovery rate (FDR).

#### Sensitivity analyses

To consider the influence of data coverage, we repeated our main analyses in restricted subsets of the cohort, including only individuals with complete or relatively complete data coverage (≥95%) during the time each individual was included in the study during childhood (ages 5-12) or adolescence (ages 12-18).

## Results

The sample included 13,593 individuals (N=2,680(19.7%) females) with ADHD and 578,793individuals (N=286,734(49.5%) females) without ADHD. Of the sample, 89.9% met inclusion criteria for analyses of childhood (ages 5–11) antecedents, including 12,425 individuals (N=2,448(19.7%) females) with ADHD and 520,097 (257,612(49.5%) females) without ADHD, while 94.9% met inclusion criteria for analyses of adolescent/adult (ages 12– 25) variables. This included 13,248 individuals (2,618(19.8%) females) with ADHD and 548,922 (272,093(49.6%) females) without ADHD. **Table S1** presents descriptive statistics for childhood antecedents and adolescent/adult outcomes, split by ADHD diagnosis and sex.

### Childhood (ages 5–11) antecedents of later ADHD diagnosis

#### Earlier vs later diagnosis

After accounting for multiple testing, the following childhood antecedents were more common in earlier (N=1,366) compared to later (N=1,082) diagnosed females: autism, learning difficulties, conduct disorder, anti-anxiety medication prescriptions, maternal depression, KS1 and KS2 failure, contacts with healthcare services (GP, outpatients, and inpatients), and socioeconomic deprivation. Detailed results are presented in **Figures 1-2 & Table 1**. The following childhood antecedents showed no evidence of strong association with age at ADHD diagnosis: anxiety, maternal ADHD, school absences, ethnic minority status, being care-experienced, and emergency care visits.

**Table 1:** Sex-stratified and interaction analyses of childhood antecedents (ages 5-11) and ADHD diagnosis timing (earlier vs later)

| Variable | Females |  |  | Males |  |  | Sex-by-predictor interaction analysis |  |
| --- | --- | --- | --- | --- | --- | --- | --- | --- |
|  | N | OR (95% CIs) | p<sub>FDR</sub> | N | OR (95% CIs) | p<sub>FDR</sub> | OR (95% CIs) | p<sub>FDR</sub> |
| Autism | 2448 | 0.20 (0.12-0.32) | **1.60E-10** | 9977 | 0.33 (0.27-0.40) | **3.26E-26** | 0.62 (0.37-1.05) | 0.22 |
| Learning difficulties | 2448 | 0.29 (0.18-0.46) | **1.92E-07** | 9977 | 0.47 (0.37-0.59) | **1.05E-09** | 0.63 (0.38-1.05) | 0.22 |
| Conduct disorder | 2448 | 0.21 (0.14-0.32) | **2.25E-13** | 9977 | 0.44 (0.37-0.52) | **1.02E-20** | 0.49 (0.31-0.76) | **0.025** |
| Anxiety | 2448 | 0.86 (0.52-1.43) | 0.64 | 9977 | 0.75 (0.55-1.02) | 0.076 | 1.18 (0.65-2.13) | 0.81 |
| Anti-anxiety medication | 2448 | 0.18 (0.12-0.27) | **1.31E-14** | 9977 | 0.18 (0.14-0.23) | **8.22E-41** | 1.00 (0.61-1.62) | 0.99 |
| Maternal ADHD* | 1965 | 1.71 (1.03-2.84) | 0.055 | 8263 | 0.70 (0.51-0.96) | **0.038** | 2.39 (1.34-4.24) | **0.026** |
| Maternal depression | 1965 | 0.79 (0.65-0.95) | **0.021** | 8263 | 0.79 (0.72-0.87) | **5.97E-06** | 1.01 (0.82-1.25) | 0.99 |
| Ethnic minority* | 1860 | 1.45 (0.83-2.56) | 0.24 | 7446 | 1.24 (0.91-1.68) | 0.20 | 1.18 (0.62-2.24) | 0.81 |
| Care-experience* | 2448 | 0.95 (0.72-1.27) | 0.75 | 9977 | 0.96 (0.81-1.13) | 0.61 | 1.03 (0.74-1.42) | 0.99 |
| Key stage 1 failure | 1246 | 0.47 (0.37-0.59) | **1.02E-09** | 4893 | 0.47 (0.41-0.53) | **1.30E-28** | 0.99 (0.76-1.31) | 0.99 |
| Key stage 2 failure | 1688 | 0.49 (0.40-0.60) | **3.55E-11** | 6660 | 0.58 (0.52-0.65) | **1.46E-21** | 0.87 (0.70-1.10) | 0.46 |
| School absences | 1552 | 0.96 (0.78-1.19) | 0.75 | 6165 | 0.88 (0.78-0.99) | **0.039** | 1.08 (0.85-1.36) | 0.81 |
| GP contacts | 2448 | 0.86 (0.84-0.88) | **7.18E-33** | 9977 | 0.83 (0.82-0.85) | **6.14E-134** | 1.04 (1.01-1.07) | **0.047** |
| Outpatient contacts | 2448 | 0.62 (0.56-0.68) | **2.94E-21** | 9977 | 0.58 (0.54-0.61) | **4.75E-69** | 1.08 (0.97-1.20) | 0.37 |
| Inpatient contacts | 2448 | 0.66 (0.50-0.87) | **6.32E-03** | 9977 | 0.81 (0.70-0.94) | **0.011** | 0.83 (0.61-1.14) | 0.46 |
| Emergency contacts | 2448 | 1.36 (0.94-1.98) | 0.13 | 9977 | 1.64 (1.33-2.01) | **5.89E-06** | 0.91 (0.64-1.30) | 0.81 |
| WIMD** | 2433 | 0.92 (0.87-0.98) | **0.017** | 9906 | 0.99 (0.96-1.02) | 0.53 | 0.94 (0.88-1.00) | 0.22 |

\*Lifetime (not restricted to ages 5–11); \*\*At entry into study. Outcome variable is coded as 0=earlier & 1=later. FDR: false discovery rate; WIMD: Welsh Index of Multiple Deprivation (higher = greater deprivation). Bold font indicates p<sub>FDR</sub><0.05.

**Figure 1.**
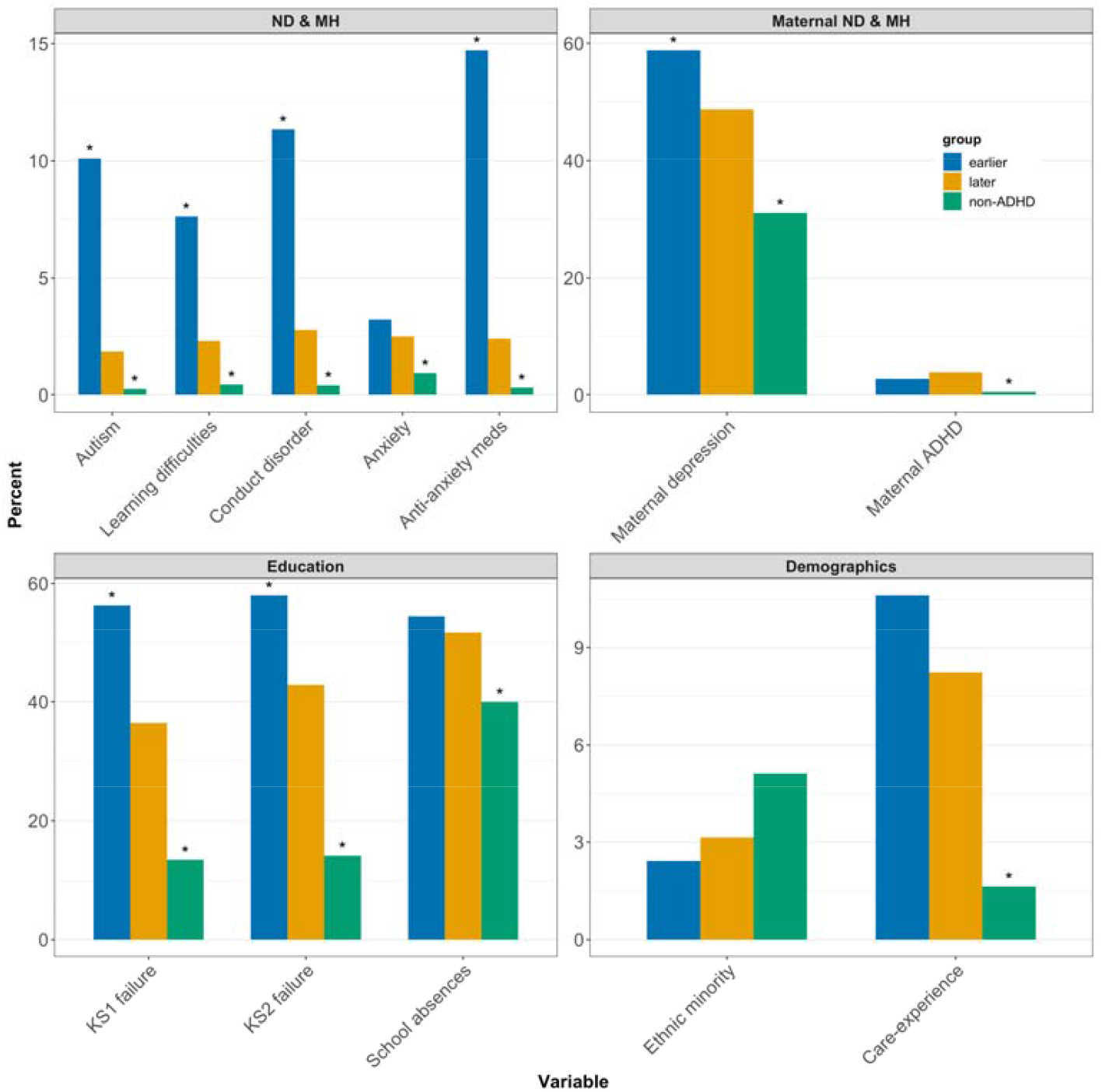
Binary childhood antecedents (ages 5-11) in females with earlier vs later vs no ADHD diagnosis. *p_FDR_<0.05. Asterisks above the earlier ADHD group indicate that those results differ from the later ADHD group. Asterisks above the non-ADHD group indicate that those results differ from the later ADHD group. ND: neurodevelopmental; MH: mental health; KS: Key Stage.

**Figure 2.**
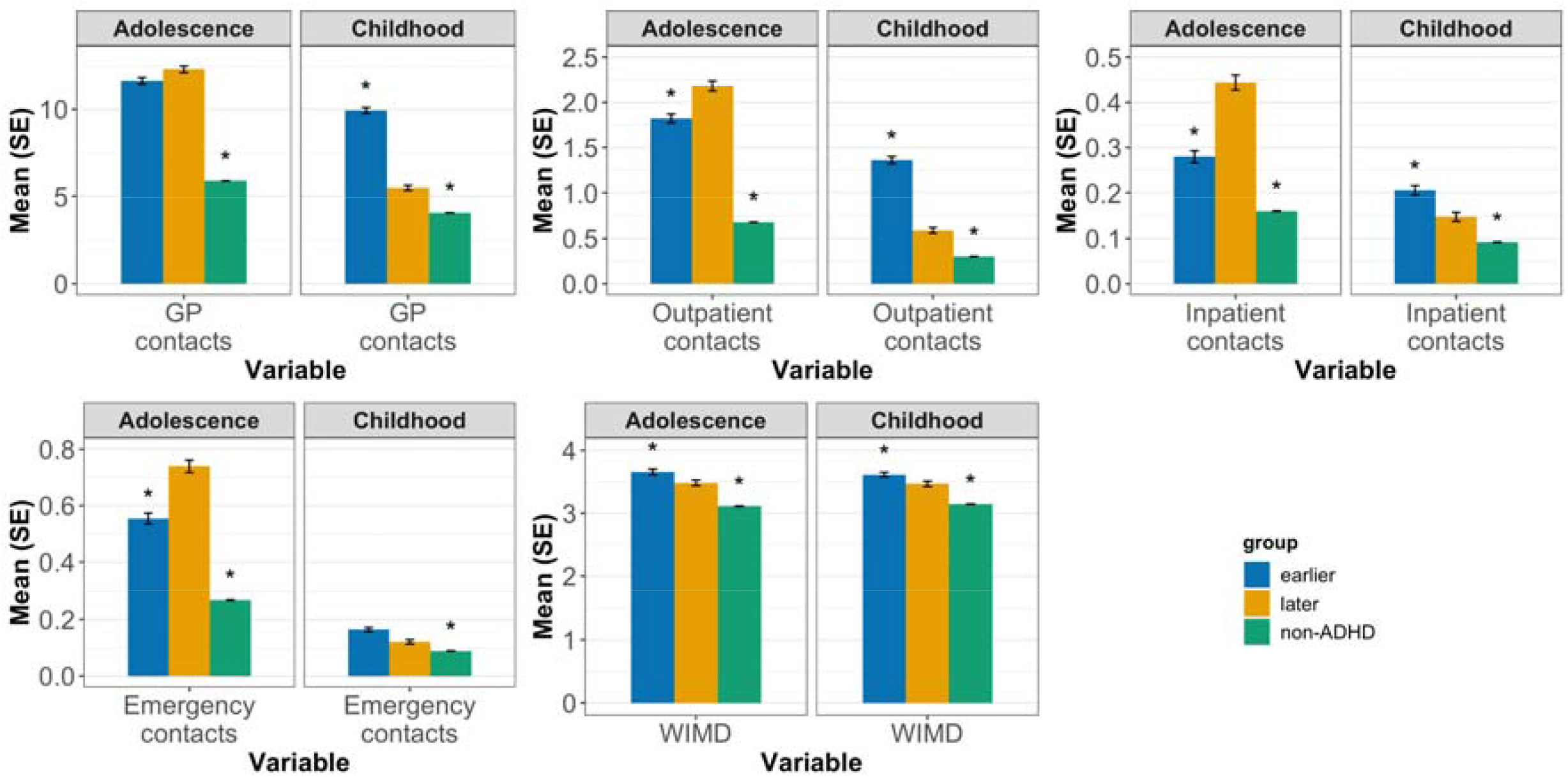
Continuous childhood antecedents and adolescent/adult outcomes in females with earlier vs later vs no ADHD diagnosis. *p_FDR_<0.05. Asterisks above the earlier ADHD group indicate that those results differ from the later ADHD group. Asterisks above the non-ADHD group indicate that those results differ from the later ADHD group. GP: General Practice (primary care); WIMD: Welsh Index of Multiple Deprivation (higher = greater deprivation).

Interaction analyses suggested a largely similar pattern of results in males (**Table 1**). The exceptions were that the association for conduct disorder was stronger in females, whereas for GP contacts this was stronger in males. There was also a sex difference for maternal ADHD; while males with earlier ADHD diagnosis were more likely to have a mother with ADHD compared to later diagnosed males, the pattern was reversed for females, albeit this was not significant. Additionally, later ADHD was associated with school absences and emergency care contacts in males, whereas socioeconomic deprivation was not associated, though the strengths of these associations did not differ by sex.

#### Later vs no diagnosis

When females with later diagnosed ADHD (after age 12), were compared with females without ADHD (N=257,612) in childhood (ages 5–11), they had higher rates of: autism, learning difficulties, conduct disorder, anxiety, anti-anxiety medication prescriptions, maternal depression, maternal ADHD, KS1 and KS2 failure, school absences, being care-experienced, more contacts with all healthcare services, and greater socioeconomic deprivation. There was no difference for ethnic minority status. Detailed results are presented in **Figures 1-2 & Table 2**.

**Table 2:** Sex-stratified and interaction analyses of childhood antecedents (ages 5-11) and later ADHD diagnosis compared to no ADHD.

| Variable | Females |  |  | Males |  |  | Sex-by-predictor interaction analysis |  |
| --- | --- | --- | --- | --- | --- | --- | --- | --- |
|  | N | OR (95% CIs) | p<sub>FDR</sub> | N | OR (95% CIs) | p<sub>FDR</sub> | OR (95% CIs) | p<sub>FDR</sub> |
| Autism | 258694 | 7.72 (4.91-12.12) | **1.2E-18** | 265633 | 3.41 (2.82-4.12) | **2.5E-36** | 2.41 (1.48-3.93) | **7.12E-03** |
| Learning difficulties | 258694 | 5.19 (3.47-7.76) | **1.5E-15** | 265633 | 3.20 (2.57-3.99) | **4.3E-25** | 1.69 (1.07-2.67) | 0.060 |
| Conduct disorder | 258694 | 6.75 (4.67-9.76) | **6.3E-24** | 265633 | 7.12 (6.08-8.35) | **1.8E-128** | 0.99 (0.66-1.47) | 0.94 |
| Anxiety | 258694 | 2.75 (1.87-4.05) | **2.8E-07** | 265633 | 2.27 (1.75-2.96) | **1.1E-09** | 1.27 (0.80-2.03) | 0.41 |
| Anti-anxiety medication | 258694 | 8.42 (5.65-12.55) | **2.6E-25** | 265633 | 5.92 (4.64-7.54) | **2.6E-46** | 1.48 (0.93-2.37) | 0.17 |
| Maternal ADHD* | 215481 | 9.02 (6.14-13.23) | **8.8E-29** | 221374 | 5.74 (4.35-7.57) | **6.3E-35** | 1.61 (1.03-2.53) | 0.081 |
| Maternal depression | 215481 | 2.13 (1.85-2.44) | **1.8E-26** | 221374 | 2.01 (1.85-2.18) | **2.3E-62** | 1.11 (0.95-1.30) | 0.25 |
| Ethnic minority* | 198511 | 0.67 (0.44-1.02) | 0.062 | 203319 | 0.60 (0.47-0.76) | **3.5E-05** | 1.11 (0.69-1.81) | 0.70 |
| Care-experience* | 258694 | 5.75 (4.59-7.19) | **5.8E-52** | 265633 | 5.46 (4.69-6.35) | **1.1E-106** | 1.12 (0.86-1.46) | 0.49 |
| Key stage 1 failure | 115318 | 3.75 (3.11-4.52) | **5.9E-43** | 118372 | 2.73 (2.44-3.06) | **1.7E-66** | 1.37 (1.10-1.70) | **0.020** |
| Key stage 2 failure | 158104 | 4.36 (3.73-5.10) | **1.2E-74** | 162098 | 3.50 (3.19-3.84) | **7.2E-152** | 1.25 (1.05-1.49) | **0.040** |
| School absences | 148507 | 1.74 (1.49-2.04) | **5.9E-12** | 151675 | 1.84 (1.67-2.03) | **5.2E-34** | 0.94 (0.78-1.12) | 0.54 |
| GP contacts | 258694 | 1.07 (1.06-1.08) | **1.5E-36** | 265633 | 1.05 (1.05-1.06) | **2.9E-61** | 1.02 (1.01-1.03) | **0.020** |
| Outpatient contacts | 258694 | 1.52 (1.45-1.60) | **1.3E-58** | 265633 | 1.44 (1.40-1.49) | **6.4E-111** | 1.09 (1.03-1.15) | **0.020** |
| Inpatient contacts | 258694 | 1.85 (1.62-2.11) | **1.6E-19** | 265633 | 1.61 (1.48-1.75) | **1.6E-27** | 1.16 (1.00-1.36) | 0.11 |
| Emergency contacts | 258694 | 3.67 (2.71-4.96) | **4.6E-17** | 265633 | 3.48 (2.93-4.15) | **1.7E-44** | 1.22 (0.93-1.60) | 0.23 |
| WIMD** | 257614 | 1.16 (1.11-1.21) | **1.2E-11** | 264539 | 1.24 (1.21-1.27) | **2.9E-57** | 0.94 (0.89-0.99) | **0.045** |

\*Lifetime (not restricted to ages 5-11); \*\*At entry into study. Outcome variable is coded as 0=non-ADHD & 1=later diagnosed ADHD. FDR: false discovery rate; WIMD: Welsh Index of Multiple Deprivation (higher = greater deprivation). Bold font indicates p<sub>FDR</sub><0.05.

The associations were stronger in females compared to males for: autism, KS1 and KS2 failure, and contacts with GP and outpatient services, but with converse findings for socioeconomic deprivation. Males with later diagnosed ADHD were less likely to be from an ethnic minority compared to males without ADHD, though this did not differ by sex. The results were similar in females and males for all other variables (**Table 2**).

### Association between ADHD diagnosis timing and adolescent/adult (ages 12–25) outcomes

#### Earlier diagnosis vs later diagnosis

Compared with earlier ADHD diagnosis (N=1,326), later diagnosis (N=1,292) in females was associated with the following outcomes during adolescence/adulthood: conduct disorder, anxiety, depression, self-harm, alcohol use, drug use, bipolar disorder, schizophrenia, anti-anxiety and antidepressant medication prescriptions, teenage pregnancy, school absences, and more outpatient, inpatient, and emergency contacts. In contrast, the following adolescent outcomes were associated with an earlier ADHD diagnosis in females: KS3 and KS4 failure, low pass at KS4, and socioeconomic deprivation. There was no evidence of association with the following adolescent outcomes: autism, learning difficulties, eating disorders, antipsychotic medication prescriptions, or GP contacts. Detailed results are presented in **Figures 2-3 & Table 3**.

**Table 3:**
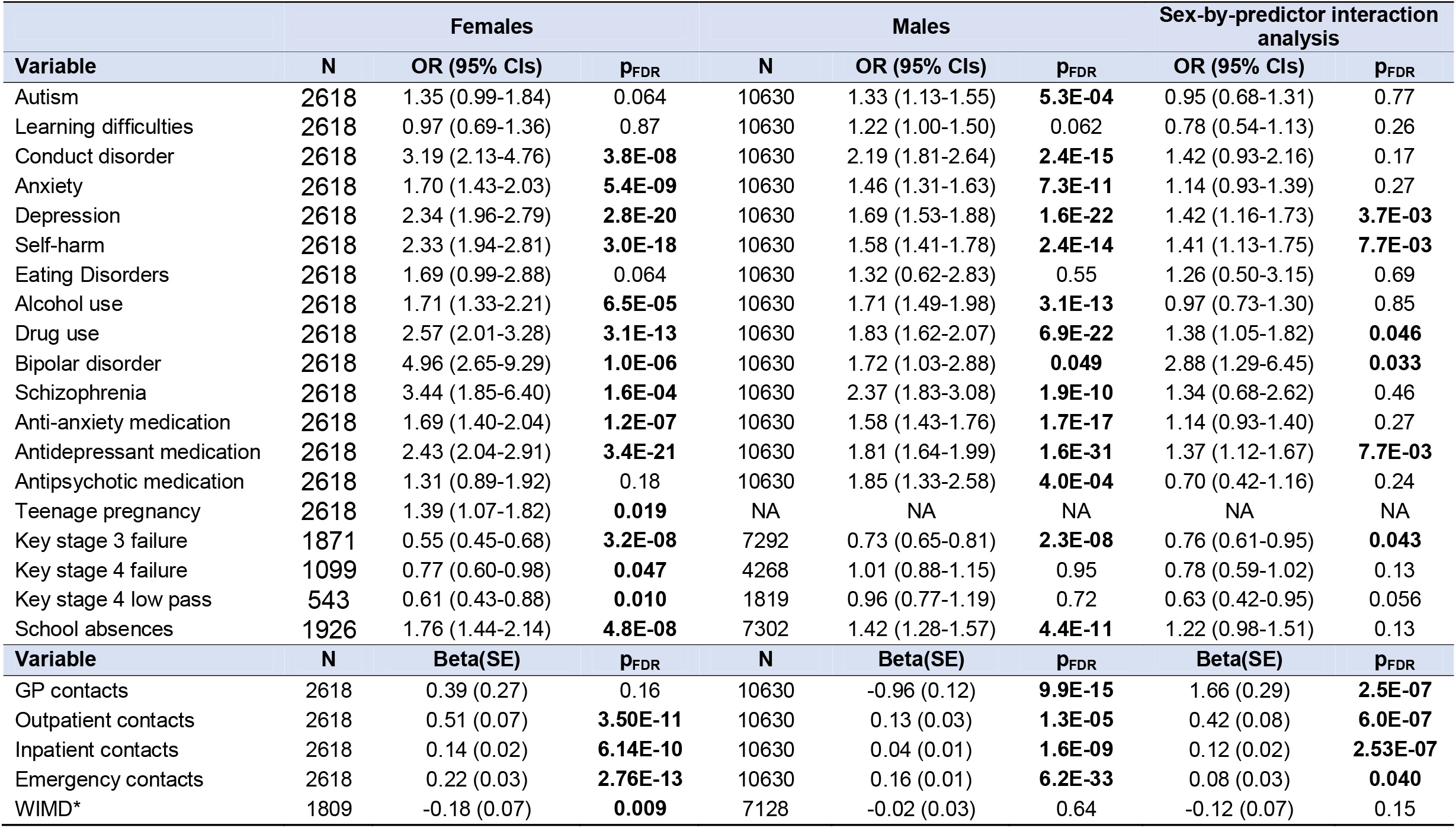
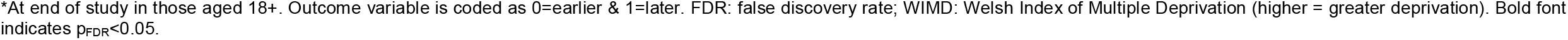
Sex-stratified and interaction analyses of ADHD diagnosis timing (earlier vs later) and adolescent/adult outcomes (ages 12-25)

|  | Females |  |  | Males |  |  | Sex-by-predictor interaction analysis |  |
| --- | --- | --- | --- | --- | --- | --- | --- | --- |
| Variable | N | OR (95% CIs) | p<sub>FDR</sub> | N | OR (95% CIs) | p<sub>FDR</sub> | OR (95% CIs) | p<sub>FDR</sub> |
| Autism | 2618 | 1.35 (0.99-1.84) | 0.064 | 10630 | 1.33 (1.13-1.55) | **5.3E-04** | 0.95 (0.68-1.31) | 0.77 |
| Learning difficulties | 2618 | 0.97 (0.69-1.36) | 0.87 | 10630 | 1.22 (1.00-1.50) | 0.062 | 0.78 (0.54-1.13) | 0.26 |
| Conduct disorder | 2618 | 3.19 (2.13-4.76) | **3.8E-08** | 10630 | 2.19 (1.81-2.64) | **2.4E-15** | 1.42 (0.93-2.16) | 0.17 |
| Anxiety | 2618 | 1.70 (1.43-2.03) | **5.4E-09** | 10630 | 1.46 (1.31-1.63) | **7.3E-11** | 1.14 (0.93-1.39) | 0.27 |
| Depression | 2618 | 2.34 (1.96-2.79) | **2.8E-20** | 10630 | 1.69 (1.53-1.88) | **1.6E-22** | 1.42 (1.16-1.73) | **3.7E-03** |
| Self-harm | 2618 | 2.33 (1.94-2.81) | **3.0E-18** | 10630 | 1.58 (1.41-1.78) | **2.4E-14** | 1.41 (1.13-1.75) | **7.7E-03** |
| Eating Disorders | 2618 | 1.69 (0.99-2.88) | 0.064 | 10630 | 1.32 (0.62-2.83) | 0.55 | 1.26 (0.50-3.15) | 0.69 |
| Alcohol use | 2618 | 1.71 (1.33-2.21) | **6.5E-05** | 10630 | 1.71 (1.49-1.98) | **3.1E-13** | 0.97 (0.73-1.30) | 0.85 |
| Drug use | 2618 | 2.57 (2.01-3.28) | **3.1E-13** | 10630 | 1.83 (1.62-2.07) | **6.9E-22** | 1.38 (1.05-1.82) | **0.046** |
| Bipolar disorder | 2618 | 4.96 (2.65-9.29) | **1.0E-06** | 10630 | 1.72 (1.03-2.88) | **0.049** | 2.88 (1.29-6.45) | **0.033** |
| Schizophrenia | 2618 | 3.44 (1.85-6.40) | **1.6E-04** | 10630 | 2.37 (1.83-3.08) | **1.9E-10** | 1.34 (0.68-2.62) | 0.46 |
| Anti-anxiety medication | 2618 | 1.69 (1.40-2.04) | **1.2E-07** | 10630 | 1.58 (1.43-1.76) | **1.7E-17** | 1.14 (0.93-1.40) | 0.27 |
| Antidepressant medication | 2618 | 2.43 (2.04-2.91) | **3.4E-21** | 10630 | 1.81 (1.64-1.99) | **1.6E-31** | 1.37 (1.12-1.67) | **7.7E-03** |
| Antipsychotic medication | 2618 | 1.31 (0.89-1.92) | 0.18 | 10630 | 1.85 (1.33-2.58) | **4.0E-04** | 0.70 (0.42-1.16) | 0.24 |
| Teenage pregnancy | 2618 | 1.39 (1.07-1.82) | **0.019** | NA | NA | NA | NA | NA |
| Key stage 3 failure | 1871 | 0.55 (0.45-0.68) | **3.2E-08** | 7292 | 0.73 (0.65-0.81) | **2.3E-08** | 0.76 (0.61-0.95) | **0.043** |
| Key stage 4 failure | 1099 | 0.77 (0.60-0.98) | **0.047** | 4268 | 1.01 (0.88-1.15) | 0.95 | 0.78 (0.59-1.02) | 0.13 |
| Key stage 4 low pass | 543 | 0.61 (0.43-0.88) | **0.010** | 1819 | 0.96 (0.77-1.19) | 0.72 | 0.63 (0.42-0.95) | 0.056 |
| School absences | 1926 | 1.76 (1.44-2.14) | **4.8E-08** | 7302 | 1.42 (1.28-1.57) | **4.4E-11** | 1.22 (0.98-1.51) | 0.13 |
| Variable | N | Beta(SE) | p<sub>FDR</sub> | N | Beta(SE) | p<sub>FDR</sub> | Beta(SE) | p<sub>FDR</sub> |
| GP contacts | 2618 | 0.39 (0.27) | 0.16 | 10630 | -0.96 (0.12) | **9.9E-15** | 1.66 (0.29) | **2.5E-07** |
| Outpatient contacts | 2618 | 0.51 (0.07) | **3.50E-11** | 10630 | 0.13 (0.03) | **1.3E-05** | 0.42 (0.08) | **6.0E-07** |
| Inpatient contacts | 2618 | 0.14 (0.02) | **6.14E-10** | 10630 | 0.04 (0.01) | **1.6E-09** | 0.12 (0.02) | **2.53E-07** |
| Emergency contacts | 2618 | 0.22 (0.03) | **2.76E-13** | 10630 | 0.16 (0.01) | **6.2E-33** | 0.08 (0.03) | **0.040** |
| WIMD* | 1809 | -0.18 (0.07) | **0.009** | 7128 | -0.02 (0.03) | 0.64 | -0.12 (0.07) | 0.15 |

\*At end of study in those aged 18+. Outcome variable is coded as 0=earlier & 1=later. FDR: false discovery rate; WIMD: Welsh Index of Multiple Deprivation (higher = greater deprivation). Bold font indicates $p_{FDR} < 0.05$ .

**Figure 3.**
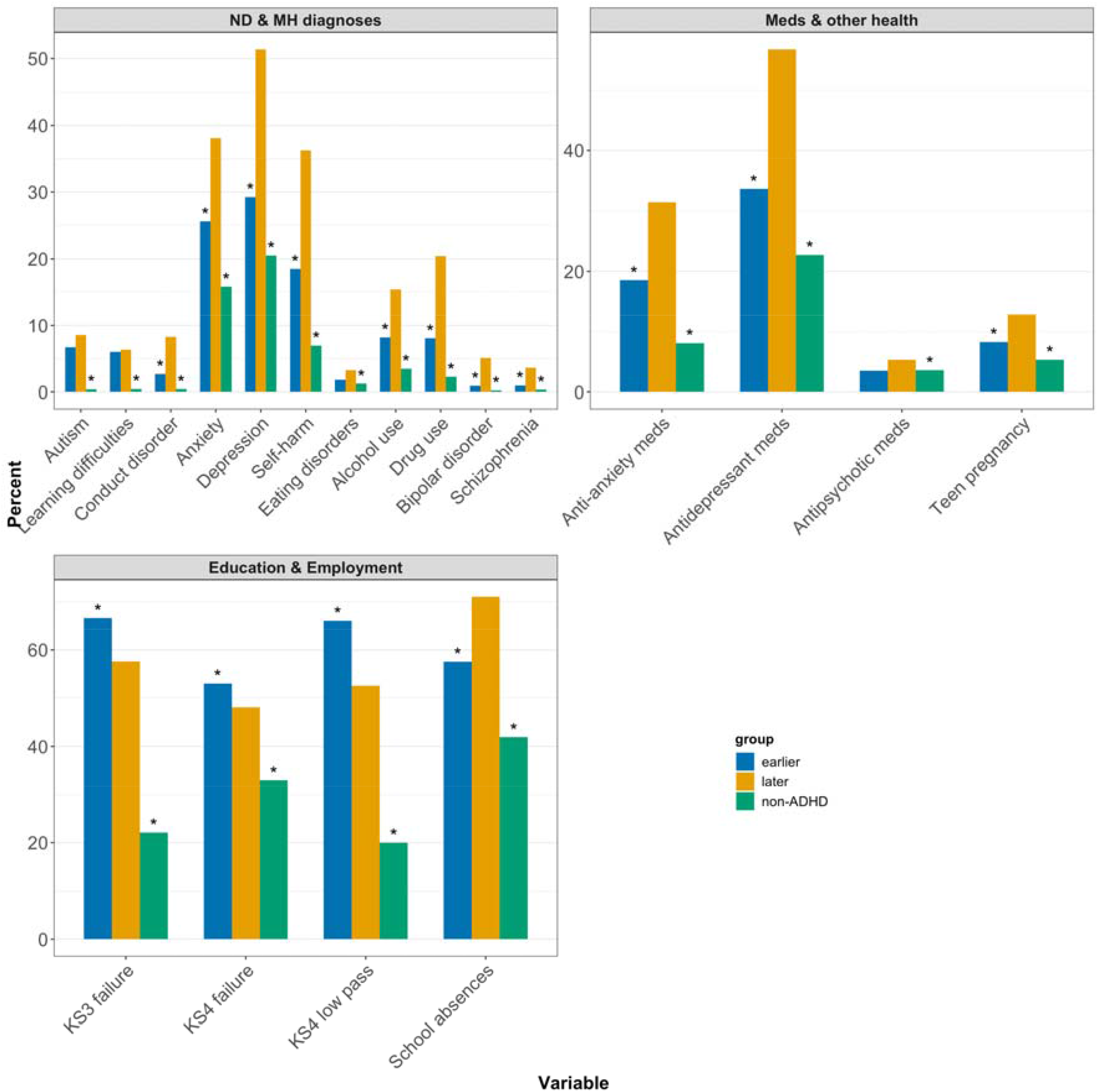
Binary adolescent/adult outcomes (ages 12-25) in females with earlier vs later vs no ADHD diagnosis. *p_FDR_<0.05. Asterisks above the earlier ADHD group indicate that those results differ from the later ADHD group. Asterisks above the non-ADHD group indicate that those results differ from the later ADHD group. ND: neurodevelopmental; MH: mental health; KS: Key Stage.

The associations were stronger in females compared to males (with the same direction of effects) for depression, self-harm, drug use, bipolar disorder, anti-depressant prescriptions, KS3 failure, and contacts with outpatient, inpatient, and emergency services. The association for GP contacts was stronger in males (earlier diagnosed males had more GP contacts), with no evidence of association in females. A few variables showed no evidence of association in males (KS4 failure and low pass) or showed evidence of association only in males (autism and antipsychotic medication prescriptions), though the interaction analyses showed no differences by sex. All other variables showed similar results in females and males (**Table 3**).

#### Later vs no diagnosis

Females with ADHD diagnosed after age 12, were more likely than females without ADHD (N=272,093) to experience the following outcomes in adolescence/adulthood: all examined conditions and prescriptions, teenage pregnancy, KS3 and KS4 failure, low pass at KS4, school absences, more contacts with all healthcare services, and higher socioeconomic deprivation. Detailed results are presented in **Figures 2-3 & Table 4**.

**Table 4:**
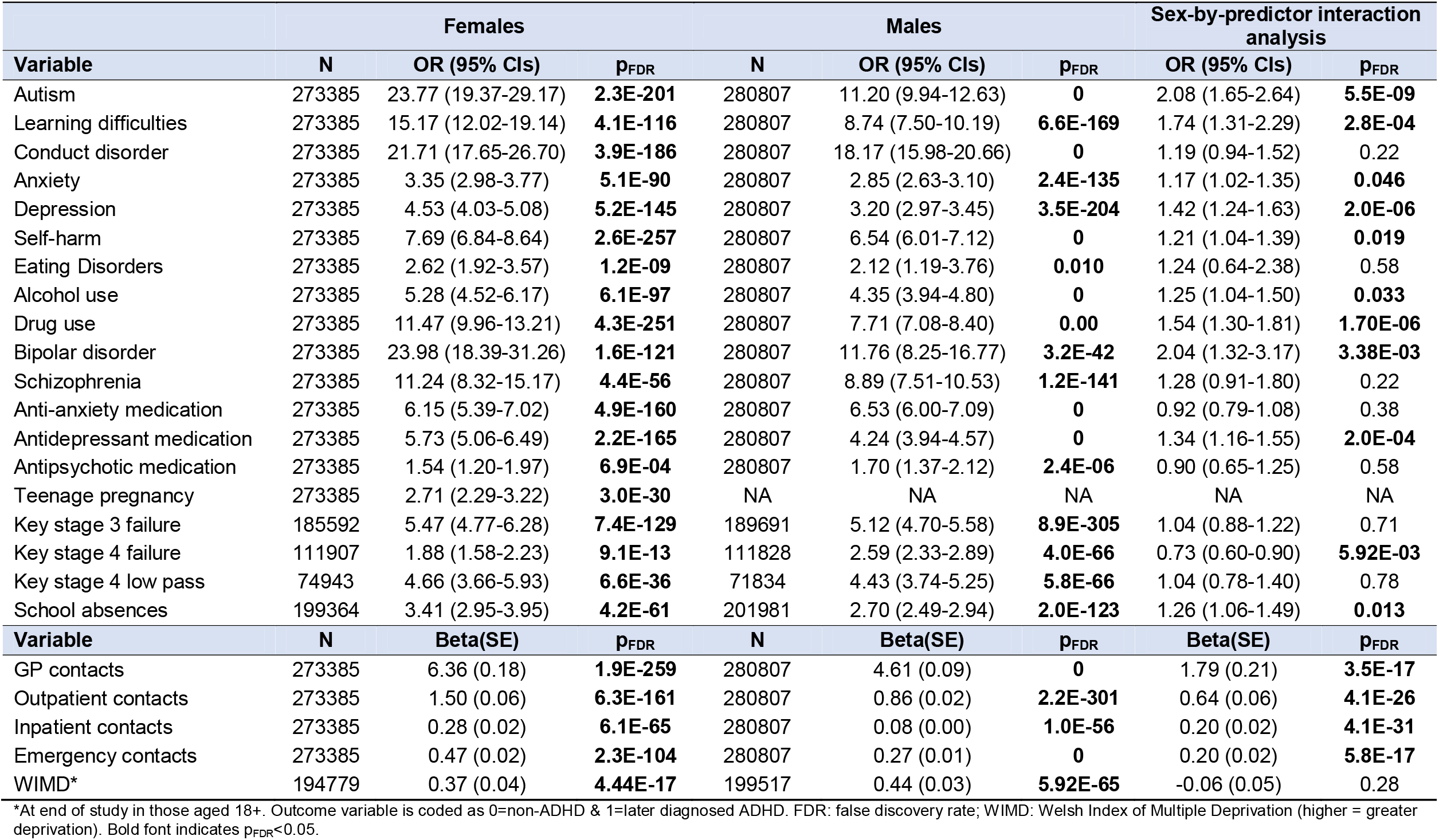
Sex-stratified and interaction analyses of later ADHD diagnosis vs no ADHD and adolescent/adult outcomes (ages 12-25)

| Variable | Females |  |  | Males |  |  | Sex-by-predictor interaction analysis |  |
| --- | --- | --- | --- | --- | --- | --- | --- | --- |
|  | N | OR (95% CIs) | p<sub>FDR</sub> | N | OR (95% CIs) | p<sub>FDR</sub> | OR (95% CIs) | p<sub>FDR</sub> |
| Autism | 273385 | 23.77 (19.37-29.17) | **2.3E-201** | 280807 | 11.20 (9.94-12.63) | **0** | 2.08 (1.65-2.64) | **5.5E-09** |
| Learning difficulties | 273385 | 15.17 (12.02-19.14) | **4.1E-116** | 280807 | 8.74 (7.50-10.19) | **6.6E-169** | 1.74 (1.31-2.29) | **2.8E-04** |
| Conduct disorder | 273385 | 21.71 (17.65-26.70) | **3.9E-186** | 280807 | 18.17 (15.98-20.66) | **0** | 1.19 (0.94-1.52) | 0.22 |
| Anxiety | 273385 | 3.35 (2.98-3.77) | **5.1E-90** | 280807 | 2.85 (2.63-3.10) | **2.4E-135** | 1.17 (1.02-1.35) | **0.046** |
| Depression | 273385 | 4.53 (4.03-5.08) | **5.2E-145** | 280807 | 3.20 (2.97-3.45) | **3.5E-204** | 1.42 (1.24-1.63) | **2.0E-06** |
| Self-harm | 273385 | 7.69 (6.84-8.64) | **2.6E-257** | 280807 | 6.54 (6.01-7.12) | **0** | 1.21 (1.04-1.39) | **0.019** |
| Eating Disorders | 273385 | 2.62 (1.92-3.57) | **1.2E-09** | 280807 | 2.12 (1.19-3.76) | **0.010** | 1.24 (0.64-2.38) | 0.58 |
| Alcohol use | 273385 | 5.28 (4.52-6.17) | **6.1E-97** | 280807 | 4.35 (3.94-4.80) | **0** | 1.25 (1.04-1.50) | **0.033** |
| Drug use | 273385 | 11.47 (9.96-13.21) | **4.3E-251** | 280807 | 7.71 (7.08-8.40) | **0.00** | 1.54 (1.30-1.81) | **1.70E-06** |
| Bipolar disorder | 273385 | 23.98 (18.39-31.26) | **1.6E-121** | 280807 | 11.76 (8.25-16.77) | **3.2E-42** | 2.04 (1.32-3.17) | **3.38E-03** |
| Schizophrenia | 273385 | 11.24 (8.32-15.17) | **4.4E-56** | 280807 | 8.89 (7.51-10.53) | **1.2E-141** | 1.28 (0.91-1.80) | 0.22 |
| Anti-anxiety medication | 273385 | 6.15 (5.39-7.02) | **4.9E-160** | 280807 | 6.53 (6.00-7.09) | **0** | 0.92 (0.79-1.08) | 0.38 |
| Antidepressant medication | 273385 | 5.73 (5.06-6.49) | **2.2E-165** | 280807 | 4.24 (3.94-4.57) | **0** | 1.34 (1.16-1.55) | **2.0E-04** |
| Antipsychotic medication | 273385 | 1.54 (1.20-1.97) | **6.9E-04** | 280807 | 1.70 (1.37-2.12) | **2.4E-06** | 0.90 (0.65-1.25) | 0.58 |
| Teenage pregnancy | 273385 | 2.71 (2.29-3.22) | **3.0E-30** | NA | NA | NA | NA | NA |
| Key stage 3 failure | 185592 | 5.47 (4.77-6.28) | **7.4E-129** | 189691 | 5.12 (4.70-5.58) | **8.9E-305** | 1.04 (0.88-1.22) | 0.71 |
| Key stage 4 failure | 111907 | 1.88 (1.58-2.23) | **9.1E-13** | 111828 | 2.59 (2.33-2.89) | **4.0E-66** | 0.73 (0.60-0.90) | **5.92E-03** |
| Key stage 4 low pass | 74943 | 4.66 (3.66-5.93) | **6.6E-36** | 71834 | 4.43 (3.74-5.25) | **5.8E-66** | 1.04 (0.78-1.40) | 0.78 |
| School absences | 199364 | 3.41 (2.95-3.95) | **4.2E-61** | 201981 | 2.70 (2.49-2.94) | **2.0E-123** | 1.26 (1.06-1.49) | **0.013** |
| Variable | N | Beta(SE) | p<sub>FDR</sub> | N | Beta(SE) | p<sub>FDR</sub> | Beta(SE) | p<sub>FDR</sub> |
| GP contacts | 273385 | 6.36 (0.18) | **1.9E-259** | 280807 | 4.61 (0.09) | **0** | 1.79 (0.21) | **3.5E-17** |
| Outpatient contacts | 273385 | 1.50 (0.06) | **6.3E-161** | 280807 | 0.86 (0.02) | **2.2E-301** | 0.64 (0.06) | **4.1E-26** |
| Inpatient contacts | 273385 | 0.28 (0.02) | **6.1E-65** | 280807 | 0.08 (0.00) | **1.0E-56** | 0.20 (0.02) | **4.1E-31** |
| Emergency contacts | 273385 | 0.47 (0.02) | **2.3E-104** | 280807 | 0.27 (0.01) | **0** | 0.20 (0.02) | **5.8E-17** |
| WIMD* | 194779 | 0.37 (0.04) | **4.44E-17** | 199517 | 0.44 (0.03) | **5.92E-65** | -0.06 (0.05) | 0.28 |

\*At end of study in those aged 18+. Outcome variable is coded as 0=non-ADHD & 1=later diagnosed ADHD. FDR: false discovery rate; WIMD: Welsh Index of Multiple Deprivation (higher = greater deprivation). Bold font indicates p<sub>FDR</sub><0.05.

The associations between later diagnoses compared to no diagnosis were stronger in females compared to males for eight of the conditions (autism, learning difficulties, anxiety, depression, self-harm, alcohol use, drug use, and bipolar disorder), antidepressant prescriptions, school absences, and all healthcare contacts. On the other hand, the association was stronger in males for KS4 failure. The results were similar in females and males for all other variables (**Table 4**).

### Sensitivity tests

We repeated analyses in subsets of individuals with relatively complete (≥95%) data coverage (50.9% at ages 5-12 and 63.3% at ages 12-18). Overall, the results were broadly similar to the primary analyses, aside from the following differences (**Tables S2-5**). Several associations of childhood antecedents no longer differed by sex (GP contacts in the analysis of earlier vs later ADHD; KS2 failure and outpatient contacts in the later vs no ADHD analysis), whereas the association was now stronger in females for autism in the analysis of earlier vs later ADHD. In the analysis of adolescent/adult outcomes, several associations were no longer significant (teenage pregnancy and KS4 failure with later compared to earlier ADHD in females; autism with later compared to earlier ADHD in males). Also, several associations no longer differed in strength by sex (antidepressant prescriptions, KS3 failure, and emergency contacts in the earlier vs later ADHD analysis; anxiety and KS4 failure in the later vs no ADHD analysis), whereas the association was now stronger in females for conduct disorder in the analysis of later vs no ADHD.

## Discussion

This study aimed to determine the childhood antecedents and adolescent/adult outcomes of later (ages 12–25) ADHD diagnosis, compared to earlier (<12) and no diagnosis in females. Overall, the results indicate that although females with a later ADHD diagnosis generally had fewer difficulties than those with an earlier diagnosis (indicating a less severe phenotype in childhood), several childhood antecedents did not differ between these groups. Compared to females without ADHD, those receiving a later diagnosis already had clear evidence of childhood mental health and educational difficulties, increased healthcare usage, and socioeconomic deprivation. Several of these antecedents were more strongly associated in females than males. Despite these childhood differences indicating a less severe phenotype, by adolescence and early adulthood, females with a later diagnosis experienced more neurodevelopmental and mental health difficulties, teenage pregnancy, and healthcare contacts, compared to those diagnosed earlier. Many of these outcomes were exacerbated in females compared to males. All investigated adverse outcomes were more likely in females with a later diagnosis than no diagnosis, with generally stronger associations in females than males. Thus, our results demonstrate that a delay in ADHD diagnosis has profound and clear consequences by adolescence and disproportionately disadvantages females.

Our results support our first hypothesis that females with a later diagnosis have clear evidence of difficulties in childhood, which precede ADHD diagnosis. They had similar difficulties to females diagnosed earlier for anxiety, school absences, and emergency department visits. Compared to females without ADHD, they had a higher likelihood of recognised neurodevelopmental and mental health problems (autism, learning difficulties, conduct disorder, anxiety, anti-anxiety medication prescriptions), maternal ADHD and depression, educational problems (exam failure and school absences), and more contacts with healthcare services. As such, the presence of these childhood difficulties and healthcare service use, known to be associated with ADHD^39,40^, can be considered as important early indicators that ADHD assessment may be warranted in primary school age females. Females with later diagnosed ADHD were also more likely to have experiences of the care system and socioeconomic deprivation compared to females without ADHD, consistent with previous research^41,42^. These demographic groups are especially vulnerable to barriers to accessing care and to their health needs being overlooked. Overall, this evidence argues against the explanation of later diagnosis being a late-emerging ADHD phenotype that only has clinically significant impact later in life^43,44^.

The generally elevated risks of childhood healthcare and educational difficulties in females with an earlier compared to a later diagnosis are consistent with previous research^45^ and indicate that the most severely impacted females are more likely to reach ADHD services in Wales. However, our study clearly shows that those with fewer but nonetheless substantive difficulties are missing out on timely ADHD recognition and support.

The results generally support our second hypothesis, that females with later diagnosed ADHD exhibit worse healthcare, educational, and socioeconomic outcomes compared to both females with an earlier diagnosis and without ADHD, albeit with some exceptions, as discussed below. Despite the fact that the earlier diagnosed group had generally more difficulties in childhood, by adolescence, the later diagnosed group had more problems, especially with regards to mental health (including conduct disorder, anxiety, depression, self-harm, alcohol use, drug use, bipolar disorder, schizophrenia, anti-anxiety and antidepressant medication prescriptions). They were also at higher risk of teenage pregnancy, school absences, and more healthcare contacts. This suggests a potential protective effect of timely diagnosis and treatment on these later outcomes.

However, the later diagnosed group were not at higher risk for all outcomes, as the earlier diagnosed group continued to have poorer educational outcomes (more exam failure and low pass at key stage 4) and greater socioeconomic deprivation. These findings may reflect that those diagnosed later could have the impact of their ADHD symptoms somewhat buffered by relative advantages in academic ability and family socioeconomic resources^43^, whereas the earlier diagnosed group have more learning difficulties and pre-existing family adversity. The groups also did not differ for several health outcomes (including adolescent neurodevelopmental conditions, eating disorders, antipsychotic medication prescriptions, or GP contacts). This suggests that even an earlier diagnosis cannot fully mitigate against the mental health risks conferred by ADHD, and that these emerging risks are not being sufficiently proactively identified in those diagnosed earlier. Whole-system care (encompassing healthcare, social care and educational services) needs to improve to address ADHD inequalities.

Females with later diagnosed ADHD had poorer outcomes than those without ADHD on every outcome examined, across mental health (including substance misuse) and education, and for teenage pregnancy. This is critically important. Educational failure and poor mental health (including substance misuse and severe and enduring mental illness such as schizophrenia and bipolar disorder) come at substantial and wide-ranging costs to individuals, their families, and the economy. The exceptionally high prevalence of depression (51.4%) anxiety (38.1%), and self-harm (36.2%) in the later diagnosed group should be noted. It is important to also recognise the impact on future generations as these adverse outcomes are likely to contribute to intergenerational transmission of adversity and inequality.

Although the focus of this paper was primarily on females with ADHD due to the historical neglect of this group (both clinically and in research studies), we also examined males. The pattern of results was very similar and therefore, many of the results discussed above are also relevant to later diagnosis of ADHD in males. Although females are more likely to be diagnosed later, there are still many males who are impacted by late diagnosis and thus experience consequences from delayed support. We found a number of key sex differences. Conduct disorder was a particularly strong indicator of earlier diagnosis in females, which has been shown before^46^. Maternal ADHD was more likely among earlier than later diagnosed males, with no difference in females. This likely reflects small numbers with limited power to detect sex differences and also, we cannot differentiate between timing of maternal and offspring diagnoses. Childhood autism, exam failure and healthcare contacts were more strongly associated with later diagnosis compared to no diagnosis in females than males, indicating that these are especially important indicators of likely ADHD in females.

A key observation in our study is that where there were sex differences in adolescent outcomes, many of the associations were stronger in females. The risk of negative outcomes in later diagnosed females compared to earlier diagnosed females was greater than seen in males for depression, self-harm, drug use, bipolar disorder, antidepressant prescriptions, exam failure, and secondary healthcare contacts. In the comparison of later diagnosed females against those without ADHD, the associations were stronger than in males for autism, learning difficulties, anxiety, depression, self-harm, alcohol use, drug use, bipolar disorder, antidepressant prescriptions, school absences, and all healthcare contacts. This demonstrates that the consequences of later diagnosis are likely to have a larger impact on females, particularly in terms of mental health problems, leading to greater use of healthcare resources.

Our study has several notable strengths; we used a nationwide register of all individuals with ADHD in Wales, with good coverage, including both primary and secondary healthcare records, as well as linked educational and demographics variables, with detailed information on age at first diagnoses. In terms of limitations, routinely collected administrative data have variable accuracy and are not designed for research purposes, which may lead to biases. In particular, our study only includes individuals with ADHD who reached clinical services and received a diagnosis and so many individuals with undiagnosed ADHD will be among the non-ADHD group, although this misclassification would serve to reduce differences between our three cohorts, suggesting that group differences are robust. Other limitations are that the population coverage was lower for some of the linked databases (e.g. educational, maternal, and ethnicity data only from 2011 via ONS), we lacked linkage to biological fathers, and we were only able to look at biological sex and not gender identity.

## Conclusion

This study has important clinical and wider societal implications, highlighting that a substantial proportion of females with ADHD (and a relatively smaller proportion of males) are not receiving a diagnosis of ADHD by age 12 in childhood, despite prominent co-occurring neurodevelopmental, mental health, educational, and general healthcare difficulties early in life. This late diagnosed group then goes on to have a variety of serious mental health problems, educational difficulties, and socioeconomic disadvantages by adolescence and early adulthood, which are exacerbated compared to those who do receive an earlier diagnosis and support by age 12. ADHD is highly prevalent and adverse outcomes are often severe and lifelong. Therefore, timely diagnosis and intervention are essential to reduce the impact on individuals with ADHD, as well as costs to primary and secondary healthcare services and economic productivity.

## Acknowledgements

This study was funded by the Welsh Government through Health and Care Research Wales via a National Institute for Health and Care Research (NIHR) Advanced Fellowship (Ref: NIHR-FS(A)-2022) and was also supported by a NARSAD Young Investigator Grant from the Brain & Behavior Research Foundation (grant no. 27879).

KS and TF are NIHR Senior Investigators. The views expressed are those of the authors and not necessarily those of the NIHR or the Department of Health and Social Care.

This work was supported by the Adolescent Mental Health Data Platform (ADP). The ADP is funded by MQ Mental Health Research Charity (Grant Reference MQBF/3 ADP). The views expressed are entirely those of the authors and should not be assumed to be the same as those of ADP or MQ Mental Health Research Charity.

This study makes use of anonymised data held in the Secure Anonymised Information Linkage (SAIL) Databank. We would like to acknowledge all the data providers who make anonymised data available for research.

The study also makes use of data provided by the Office of National Statistics (ONS) and we acknowledge that those who carried out the original collection and analysis of the ONS data bear no responsibility for the further analysis or interpretation.

This work was supported by the Wolfson Centre for Young People’s Mental Health, established with support from the Wolfson Foundation.

## Author contributions

- Joanna Martin: acquisition of funding, study design, analyses, manuscript writing & editing
- Olivier Rouquette: data curation & manuscript editing
- Kate Langley, Miriam Cooper, Kapil Sayal, Tamsin Ford, Ann John, Anita Thapar: study design, manuscript editing

## Competing interests

Miriam Cooper has spoken about the Cwm Taf Morgannwg Neurodevelopmental Service at conferences hosted by Takeda (formerly Shire) and Flynn Pharma. These talks were not related to the content of this article and MC did not receive personal payment.

Tamsin Ford’s research group received funds for research methods consultation with Place2Be, a third sector organisation providing mental health interventions and training within schools.

KL has participated on an advisory board for Medice. This was not related to the content of this article and KL did not receive personal payment.

## Data availability

Data are available through application to the SAIL databank (http://www.saildatabank.com).

## Notes

### Author Declarations

Ethical approval for this project was granted by the Information Governance Review Panel (IGRP #1335); this is an independent body consisting of government, regulatory and professional agencies.

