## Supplemental Materials for "Antecedents and outcomes of a late attention deficit hyperactivity disorder (ADHD) diagnosis in females"

### Table S1: Descriptive statistics for childhood antecedents (at ages 5–11) and adolescent/adult outcomes (ages 12–25) in females and males with early or later ADHD diagnosis and no ADHD diagnosis

|  | **Females** | | | **Males** | | |
| --- | --- | --- | --- | --- | --- | --- |
| **Variable** | **Early ADHD group** | **Later ADHD group** | **Non-ADHD group** | **Early ADHD group** | **Later ADHD group** | **Non-ADHD group** |
| **Childhood (ages 5-12)*** | **N=1,366** | **N=1,082** | **N=257,612** | **N=6,829** | **N=3,148** | **N=262,485** |
| Autism, N(%) | 138 (10.1) | 20 (1.8) | 626 (0.2) | 796 (11.7) | 116 (3.7) | 3007 (1.1) |
| Learning difficulties, N(%) | 104 (7.6) | 25 (2.3) | 1113 (0.4) | 408 (6.0) | 85 (2.7) | 2223 (0.8) |
| Conduct disorder, N(%) | 155 (11.4) | 30 (2.8) | 1020 (0.4) | 826 (12.1) | 175 (5.6) | 2099 (0.8) |
| Anxiety, N(%) | 44 (3.2) | 27 (2.5) | 2364 (0.9) | 190 (2.8) | 58 (1.8) | 2227 (0.8) |
| Anti-anxiety medication, N(%) | 201 (14.7) | 26 (2.4) | 769 (0.3) | 958 (14.0) | 72 (2.3) | 1116 (0.4) |
| Maternal ADHD*, N(%) | 30 (2.7) | 33 (3.8) | 976 (0.5) | 203 (3.6) | 57 (2.2) | 911 (0.4) |
| Maternal depression, N(%) | 649 (58.8) | 420 (48.8) | 66611 (31.0) | 3185 (55.8) | 1168 (45.7) | 67256 (30.7) |
| Ethnic minority*, N(%) | 25 (2.4) | 26 (3.1) | 10122 (5.1) | 129 (2.5) | 68 (2.9) | 10562 (5.3) |
| Care-experience*, N(%) | 145 (10.6) | 89 (8.2) | 4261 (1.7) | 602 (8.8) | 203 (6.4) | 3840 (1.5) |
| Key stage 1 failure, N(%) | 432 (56.3) | 175 (36.5) | 15365 (13.4) | 2216 (60.9) | 527 (42.0) | 24654 (21.1) |
| Key stage 2 failure, N(%) | 569 (58.0) | 303 (42.9) | 22192 (14.1) | 2760 (57.8) | 894 (47.5) | 31868 (19.9) |
| School absences, N(%) | 500 (54.5) | 328 (51.7) | 59128 (40.0) | 2619 (58.4) | 904 (53.8) | 60458 (40.3) |
| GP contacts, mean(SE) | 9.93 (0.17) | 5.47 (0.15) | 4.09 (0.01) | 9.72 (0.07) | 5.05 (0.08) | 4.08 (0.01) |
| Outpatient contacts, mean(SE) | 1.36 (0.04) | 0.59 (0.03) | 0.30 (0.00) | 1.35 (0.02) | 0.56 (0.02) | 0.34 (0.00) |
| Inpatient contacts, mean(SE) | 0.21 (0.01) | 0.15 (0.01) | 0.09 (0.00) | 0.18 (0.00) | 0.16 (0.01) | 0.11 (0.00) |
| Emergency contacts, mean(SE) | 0.16 (0.01) | 0.12 (0.01) | 0.09 (0.00) | 0.17 (0.00) | 0.12 (0.00) | 0.10 (0.00) |
| WIMD**, mean(SE) | 3.62 (0.04) | 3.47 (0.04) | 3.15 (0.00) | 3.60 (0.02) | 3.58 (0.02) | 3.14 (0.00) |
| **Adolescence (ages 12-18)*** | **N=1,326** | **N=1,292** | **N=272,093** | **N=6,652** | **N=3,978** | **N=276,829** |
| Autism, N(%) | 89 (6.7) | 111 (8.6) | 1080 (0.4) | 414 (6.2) | 332 (8.3) | 2278 (0.8) |
| Learning difficulties, N(%) | 80 (6.0) | 82 (6.3) | 1208 (0.4) | 242 (3.6) | 193 (4.9) | 1595 (0.6) |
| Conduct disorder, N(%) | 36 (2.7) | 107 (8.3) | 1123 (0.4) | 240 (3.6) | 314 (7.9) | 1292 (0.5) |
| Anxiety, N(%) | 339 (25.6) | 492 (38.1) | 43037 (15.8) | 870 (13.1) | 745 (18.7) | 21290 (7.7) |
| Depression, N(%) | 388 (29.3) | 664 (51.4) | 55851 (20.5) | 1080 (16.2) | 1049 (26.4) | 29388 (10.6) |
| Self-harm, N(%) | 245 (18.5) | 468 (36.2) | 19016 (7.0) | 755 (11.4) | 740 (18.6) | 9622 (3.5) |
| Eating disorders, N(%) | 24 (1.8) | 42 (3.3) | 3395 (1.2) | 14 (0.2) | 12 (0.3) | 396 (0.1) |
| Alcohol use, N(%) | 109 (8.2) | 199 (15.4) | 9469 (3.5) | 433 (6.5) | 510 (12.8) | 9371 (3.4) |
| Drug use, N(%) | 107 (8.1) | 264 (20.4) | 6174 (2.3) | 652 (9.8) | 748 (18.8) | 8494 (3.1) |
| Bipolar disorder, N(%) | 12 (0.9) | 66 (5.1) | 624 (0.2) | 29 (0.4) | 36 (0.9) | 213 (0.1) |
| Schizophrenia, N(%) | 13 (1.0) | 47 (3.6) | 917 (0.3) | 96 (1.4) | 159 (4.0) | 1293 (0.5) |
| Anti-anxiety medication, N(%) | 246 (18.6) | 406 (31.4) | 22091 (8.1) | 993 (14.9) | 942 (23.7) | 13740 (5.0) |
| Antidepressant medication, N(%) | 446 (33.6) | 733 (56.7) | 61888 (22.8) | 1377 (20.7) | 1338 (33.6) | 33622 (12.2) |
| Antipsychotic medication, N(%) | 47 (3.5) | 69 (5.3) | 9772 (3.6) | 65 (1.0) | 83 (2.1) | 3399 (1.2) |
| Teenage pregnancy, N(%) | 110 (8.3) | 166 (12.9) | 14524 (5.3) | NA | NA | NA |
| Key stage 3 failure, N(%) | 630 (66.6) | 533 (57.6) | 40806 (22.1) | 3280 (69.6) | 1720 (66.7) | 55446 (29.6) |
| Key stage 4 failure, N(%) | 299 (53.0) | 257 (48.0) | 36707 (33.0) | 1622 (57.1) | 827 (57.9) | 39167 (35.5) |
| Key stage 4 low pass, N(%) | 175 (66.0) | 146 (52.5) | 14926 (20.0) | 821 (67.4) | 393 (65.4) | 21379 (30.0) |
| School absences, N(%) | 560 (57.6) | 677 (71.0) | 83193 (41.9) | 2539 (54.0) | 1640 (63.1) | 77369 (38.8) |
| GP contacts, mean(SE) | 11.64 (0.19) | 12.30 (0.18) | 5.88 (0.01) | 9.16 (0.08) | 8.25 (0.09) | 3.64 (0.01) |
| Outpatient contacts, mean(SE) | 1.82 (0.05) | 2.18 (0.05) | 0.68 (0.00) | 1.44 (0.02) | 1.36 (0.02) | 0.52 (0.00) |
| Inpatient contacts, mean(SE) | 0.28 (0.01) | 0.44 (0.02) | 0.16 (0.00) | 0.12 (0.00) | 0.16 (0.00) | 0.08 (0.00) |
| Emergency contacts, mean(SE) | 0.55 (0.02) | 0.74 (0.02) | 0.27 (0.00) | 0.46 (0.01) | 0.56 (0.01) | 0.30 (0.00) |
| WIMD***, mean(SE) | 3.66 (0.05) | 3.48 (0.04) | 3.11 (0.00) | 3.57 (0.02) | 3.52 (0.03) | 3.08 (0.00) |

*Lifetime (not restricted to ages 5-11); **At entry into study; ***At end of study in those aged 18+. WIMD: Welsh Index of Multiple Deprivation (higher = greater deprivation).

### Table S2: Sex-stratified and interaction analyses of childhood antecedents (ages 5-11) and ADHD diagnosis timing (earlier vs later) – sensitivity test using data with nearly complete (>95%) coverage

|  | **Females** | | | **Males** | | | **Sex-by-predictor interaction analysis** | |
| --- | --- | --- | --- | --- | --- | --- | --- | --- |
| **Variable** | **N** | **OR (95% CIs)** | **p** | **N** | **OR (95% CIs)** | **p** | **OR (95% CIs)** | **p** |
| Autism | 1632 | 0.18 (0.10-0.31) | 1.22E-09 | 6407 | 0.34 (0.27-0.43) | 8.17E-20 | 0.53 (0.29-0.96) | 0.037 |
| Learning difficulties | 1632 | 0.30 (0.17-0.51) | 1.46E-05 | 6407 | 0.47 (0.35-0.62) | 8.31E-08 | 0.65 (0.35-1.19) | 0.16 |
| Conduct disorder | 1632 | 0.22 (0.14-0.36) | 1.63E-09 | 6407 | 0.45 (0.36-0.55) | 6.93E-14 | 0.51 (0.30-0.87) | 0.013 |
| Anxiety | 1632 | 0.86 (0.49-1.51) | 0.59 | 6407 | 0.93 (0.66-1.30) | 0.660 | 0.91 (0.47-1.76) | 0.79 |
| Anti-anxiety medication | 1632 | 0.14 (0.09-0.24) | 1.31E-13 | 6407 | 0.18 (0.14-0.24) | 2.90E-32 | 0.76 (0.42-1.36) | 0.35 |
| Maternal ADHD* | 1383 | 1.40 (0.80-2.46) | 0.245 | 5590 | 0.63 (0.43-0.92) | 0.016 | 2.07 (1.08-3.99) | 0.029 |
| Maternal depression | 1383 | 0.79 (0.63-0.98) | 0.036 | 5590 | 0.79 (0.70-0.89) | 1.70E-04 | 0.96 (0.75-1.23) | 0.75 |
| Ethnic minority* | 1294 | 0.99 (0.51-1.94) | 0.98 | 5068 | 1.06 (0.72-1.57) | 0.75 | 0.91 (0.42-1.96) | 0.81 |
| Care-experience* | 1632 | 0.96 (0.71-1.30) | 0.79 | 6407 | 0.96 (0.80-1.16) | 0.70 | 0.96 (0.67-1.36) | 0.81 |
| Key stage 1 failure | 1124 | 0.49 (0.38-0.63) | 2.49E-08 | 4374 | 0.47 (0.41-0.55) | 1.26E-25 | 1.03 (0.77-1.37) | 0.84 |
| Key stage 2 failure | 1467 | 0.49 (0.39-0.61) | 1.78E-10 | 5679 | 0.59 (0.52-0.67) | 1.70E-17 | 0.85 (0.66-1.08) | 0.18 |
| School absences | 1352 | 0.85 (0.68-1.07) | 0.16 | 5277 | 0.86 (0.76-0.98) | 0.019 | 0.98 (0.76-1.26) | 0.87 |
| GP contacts | 1632 | 0.88 (0.85-0.90) | 8.51E-21 | 6407 | 0.85 (0.84-0.87) | 5.39E-74 | 1.03 (1.00-1.07) | 0.06 |
| Outpatient contacts | 1632 | 0.59 (0.53-0.66) | 3.67E-20 | 6407 | 0.57 (0.53-0.61) | 1.89E-56 | 1.02 (0.90-1.16) | 0.79 |
| Inpatient contacts | 1632 | 0.55 (0.38-0.79) | 1.27E-03 | 6407 | 0.69 (0.55-0.86) | 1.29E-03 | 0.80 (0.52-1.23) | 0.31 |
| Emergency contacts | 1632 | 1.40 (0.93-2.10) | 0.11 | 6407 | 1.45 (1.15-1.84) | 1.74E-03 | 0.84 (0.56-1.25) | 0.38 |
| WIMD** | 1622 | 0.91 (0.84-0.97) | 7.28E-03 | 6371 | 0.98 (0.95-1.03) | 0.46 | 0.92 (0.85-1.00) | 0.052 |

*Lifetime (not restricted to ages 5–11); **At entry into study. Outcome variable is coded as 0=earlier & 1=later. WIMD: Welsh Index of Multiple Deprivation (higher = greater deprivation).

### Table S3: Sex-stratified and interaction analyses of childhood antecedents (ages 5-11) and later ADHD diagnosis compared to no ADHD – sensitivity test using data with nearly complete (>95%) coverage

|  | **Females** | | | **Males** | | | **Sex-by-predictor interaction analysis** | |
| --- | --- | --- | --- | --- | --- | --- | --- | --- |
| **Variable** | **N** | **OR (95% CIs)** | **p** | **N** | **OR (95% CIs)** | **p** | **OR (95% CIs)** | **p** |
| Autism | 145726 | 7.13 (4.23-12.01) | 1.6E-13 | 150142 | 3.24 (2.60-4.04) | 8.7E-26 | 2.18 (1.24-3.83) | 6.92E-03 |
| Learning difficulties | 145726 | 4.76 (2.92-7.75) | 3.6E-10 | 150142 | 3.25 (2.51-4.22) | 6.8E-19 | 1.46 (0.84-2.54) | 0.18 |
| Conduct disorder | 145726 | 6.48 (4.17-10.06) | 9.7E-17 | 150142 | 6.79 (5.57-8.28) | 1.1E-79 | 0.96 (0.59-1.55) | 0.85 |
| Anxiety | 145726 | 2.67 (1.70-4.18) | 1.8E-05 | 150142 | 2.46 (1.83-3.32) | 2.7E-09 | 1.07 (0.62-1.82) | 0.82 |
| Anti-anxiety medication | 145726 | 7.19 (4.40-11.74) | 3.4E-15 | 150142 | 6.40 (4.84-8.47) | 8.2E-39 | 1.10 (0.63-1.94) | 0.73 |
| Maternal ADHD* | 129250 | 7.78 (5.04-12.01) | 2.2E-20 | 133295 | 4.90 (3.48-6.89) | 7.2E-20 | 1.56 (0.91-2.66) | 0.10 |
| Maternal depression | 129250 | 2.22 (1.88-2.63) | 8.3E-21 | 133295 | 1.99 (1.80-2.20) | 1.3E-39 | 1.10 (0.91-1.34) | 0.33 |
| Ethnic minority* | 122738 | 0.59 (0.35-1.00) | 0.051 | 126058 | 0.59 (0.43-0.81) | 1.1E-03 | 0.97 (0.52-1.81) | 0.93 |
| Care-experience* | 145726 | 6.13 (4.80-7.81) | 2.4E-48 | 150142 | 5.57 (4.71-6.57) | 4.5E-91 | 1.07 (0.80-1.43) | 0.64 |
| Key stage 1 failure | 102267 | 3.86 (3.17-4.70) | 3.2E-41 | 105163 | 2.83 (2.51-3.19) | 6.8E-65 | 1.37 (1.09-1.73) | 6.63E-03 |
| Key stage 2 failure | 134303 | 4.24 (3.57-5.04) | 1.9E-60 | 137828 | 3.71 (3.35-4.11) | 9.2E-140 | 1.18 (0.98-1.43) | 0.083 |
| School absences | 124623 | 1.62 (1.37-1.92) | 1.4E-08 | 127514 | 1.81 (1.63-2.01) | 5.6E-28 | 0.89 (0.73-1.08) | 0.24 |
| GP contacts | 145726 | 1.07 (1.06-1.08) | 1.3E-26 | 150142 | 1.05 (1.04-1.06) | 5.3E-35 | 1.02 (1.00-1.03) | 0.019 |
| Outpatient contacts | 145726 | 1.50 (1.42-1.59) | 1.6E-45 | 150142 | 1.43 (1.38-1.49) | 3.3E-84 | 1.04 (0.97-1.11) | 0.23 |
| Inpatient contacts | 145726 | 1.82 (1.54-2.16) | 3.5E-12 | 150142 | 1.55 (1.38-1.75) | 4.5E-13 | 1.17 (0.95-1.44) | 0.13 |
| Emergency contacts | 145726 | 3.99 (2.86-5.57) | 3.5E-16 | 150142 | 3.62 (2.97-4.41) | 2.5E-37 | 0.98 (0.72-1.35) | 0.92 |
| WIMD** | 145084 | 1.16 (1.09-1.22) | 2.5E-07 | 149482 | 1.26 (1.22-1.31) | 9.6E-38 | 0.92 (0.86-0.98) | 0.013 |

*Lifetime (not restricted to ages 5-12); **At entry into study. Outcome variable is coded as 0=non-ADHD & 1=later diagnosed ADHD. WIMD: Welsh Index of Multiple Deprivation (higher = greater deprivation).

### Table S4: Sex-stratified and interaction analyses of ADHD diagnosis timing (earlier vs later) and adolescent/adult outcomes (ages 12-25) – sensitivity test using data with nearly complete (>95%) coverage

|  | **Females** | | | **Males** | | | **Sex-by-predictor interaction analysis** | |
| --- | --- | --- | --- | --- | --- | --- | --- | --- |
| **Variable** | **N** | **OR (95% CIs)** | **p** | **N** | **OR (95% CIs)** | **p** | **OR (95% CIs)** | **p** |
| Autism | 1730 | 1.15 (0.81-1.63) | 0.429 | 6853 | 1.15 (0.96-1.38) | 0.14 | 1.00 (0.68-1.47) | 0.99 |
| Learning difficulties | 1730 | 0.76 (0.53-1.09) | 0.13 | 6853 | 1.20 (0.96-1.51) | 0.116 | 0.63 (0.41-0.96) | 0.03 |
| Conduct disorder | 1730 | 2.59 (1.66-4.02) | 2.5E-05 | 6853 | 1.71 (1.38-2.13) | 1.2E-06 | 1.47 (0.90-2.39) | 0.12 |
| Anxiety | 1730 | 1.45 (1.19-1.76) | 2.3E-04 | 6853 | 1.35 (1.19-1.53) | 1.9E-06 | 1.04 (0.82-1.30) | 0.76 |
| Depression | 1730 | 1.94 (1.60-2.36) | 3.2E-11 | 6853 | 1.54 (1.38-1.73) | 1.6E-14 | 1.27 (1.01-1.59) | 3.8E-02 |
| Self-harm | 1730 | 2.10 (1.70-2.59) | 3.3E-12 | 6853 | 1.47 (1.30-1.67) | 1.9E-09 | 1.41 (1.10-1.79) | 6.3E-03 |
| Eating Disorders | 1730 | 1.61 (0.91-2.83) | 0.099 | 6853 | 0.00 (0.00-0.00) | 0.52 | 1.32 (0.47-3.67) | 0.60 |
| Alcohol use | 1730 | 1.60 (1.23-2.10) | 5.8E-04 | 6853 | 1.61 (1.39-1.87) | 4.6E-10 | 0.97 (0.71-1.32) | 0.85 |
| Drug use | 1730 | 2.53 (1.94-3.29) | 6.0E-12 | 6853 | 1.71 (1.50-1.94) | 3.8E-16 | 1.48 (1.10-1.99) | 0.010 |
| Bipolar disorder | 1730 | 3.99 (2.12-7.53) | 1.9E-05 | 6853 | 1.65 (0.98-2.78) | 0.058 | 2.43 (1.07-5.53) | 0.035 |
| Schizophrenia | 1730 | 3.03 (1.62-5.67) | 5.1E-04 | 6853 | 2.30 (1.76-3.01) | 1.5E-09 | 1.27 (0.64-2.53) | 0.49 |
| Anti-anxiety medication | 1730 | 1.66 (1.34-2.06) | 3.9E-06 | 6853 | 1.43 (1.27-1.61) | 3.3E-09 | 1.18 (0.93-1.50) | 0.18 |
| Antidepressant medication | 1730 | 2.10 (1.72-2.57) | 6.2E-13 | 6853 | 1.67 (1.51-1.86) | 5.5E-22 | 1.25 (1.00-1.56) | 0.05 |
| Antipsychotic medication | 1730 | 1.06 (0.71-1.60) | 0.76 | 6853 | 1.79 (1.27-2.53) | 8.5E-04 | 0.58 (0.34-0.98) | 0.04 |
| Teenage pregnancy | 1730 | 1.19 (0.91-1.57) | 0.208 | NA | NA | NA | NA | NA |
| Key stage 3 failure | 1318 | 0.52 (0.41-0.67) | 2.7E-07 | 5061 | 0.70 (0.61-0.80) | 1.9E-07 | 0.77 (0.58-1.01) | 0.059 |
| Key stage 4 failure | 793 | 0.84 (0.62-1.12) | 0.234 | 2922 | 1.03 (0.87-1.22) | 0.71 | 0.81 (0.58-1.14) | 0.235 |
| Key stage 4 low pass | 403 | 0.62 (0.41-0.94) | 0.023 | 1267 | 0.85 (0.66-1.10) | 0.23 | 0.73 (0.45-1.18) | 0.195 |
| School absences | 1261 | 1.48 (1.16-1.88) | 1.6E-03 | 4685 | 1.33 (1.17-1.50) | 9.5E-06 | 1.11 (0.85-1.46) | 0.440 |
| **Variable** | **N** | **Beta(SE)** | **p** | **N** | **Beta(SE)** | **p** | **Beta(SE)** | **p** |
| GP contacts | 1730 | 0.32 (0.00) | 0.44 | 6853 | 0.14 (0.00) | 2.6E-12 | 1.30 (0.35) | 1.7E-04 |
| Outpatient contacts | 1730 | 0.08 (0.00) | 7.42E-08 | 6853 | 0.03 (0.00) | 6.1E-05 | 0.32 (0.09) | 2.4E-04 |
| Inpatient contacts | 1730 | 0.03 (0.00) | 2.06E-06 | 6853 | 0.01 (0.00) | 3.4E-07 | 0.09 (0.03) | 5.9E-04 |
| Emergency contacts | 1730 | 0.03 (0.00) | 4.13E-07 | 6853 | 0.01 (0.00) | 7.9E-17 | 0.05 (0.04) | 0.154 |
| WIMD* | 1680 | 0.07 (0.00) | 0.013 | 6664 | 0.04 (0.00) | 0.79 | -0.14 (0.07) | 0.06 |

*At end of study in those aged 18+. WIMD: Welsh Index of Multiple Deprivation (higher = greater deprivation). Outcome variable is coded as 0=earlier & 1=later.

### Table S5: Sex-stratified and interaction analyses of later ADHD diagnosis compared to no ADHD and adolescent/adult outcomes (at ages 12-25) – sensitivity test using data with nearly complete (>95%) coverage

|  | **Females** | | | **Males** | | | | **Sex-by-predictor interaction analysis** | |
| --- | --- | --- | --- | --- | --- | --- | --- | --- | --- |
| **Variable** | **N** | **OR (95% CIs)** | **p** | **N** | **OR (95% CIs)** | **p** | **OR (95% CIs)** | | **p** |
| Autism | 182559 | 22.30 (17.45-28.50) | 5.4E-136 | 187123 | 10.39 (8.96-12.06) | 0 | 2.17 (1.63-2.88) | | 9.8E-08 |
| Learning difficulties | 182559 | 13.31 (10.14-17.48) | 1.7E-77 | 187123 | 8.35 (6.98-9.98) | 9.8E-120 | 1.60 (1.16-2.22) | | 4.5E-03 |
| Conduct disorder | 182559 | 20.57 (16.17-26.17) | 5.3E-134 | 187123 | 14.91 (12.72-17.47) | 0 | 1.38 (1.03-1.84) | | 0.029 |
| Anxiety | 182559 | 3.10 (2.71-3.55) | 5.0E-62 | 187123 | 2.74 (2.50-3.01) | 4.6E-99 | 1.14 (0.97-1.34) | | 0.121 |
| Depression | 182559 | 4.43 (3.86-5.07) | 1.1E-101 | 187123 | 3.13 (2.88-3.40) | 1.8E-158 | 1.41 (1.20-1.65) | | 2.6E-05 |
| Self-harm | 182559 | 7.44 (6.51-8.51) | 1.3E-188 | 187123 | 6.28 (5.71-6.91) | 0 | 1.21 (1.03-1.43) | | 0.024 |
| Eating Disorders | 182559 | 2.61 (1.85-3.66) | 3.5E-08 | 187123 | 0.00 (0.00-0.00) | 0.045 | 1.32 (0.63-2.79) | | 0.46 |
| Alcohol use | 182559 | 5.18 (4.37-6.13) | 1.5E-80 | 187123 | 4.14 (3.72-4.61) | 0 | 1.29 (1.05-1.57) | | 0.015 |
| Drug use | 182559 | 11.70 (10.02-13.67) | 2.5E-211 | 187123 | 7.40 (6.74-8.14) | 0 | 1.62 (1.35-1.94) | | 2.60E-07 |
| Bipolar disorder | 182559 | 20.19 (15.03-27.13) | 1.7E-88 | 187123 | 12.38 (8.49-18.04) | 3.9E-39 | 1.64 (1.02-2.64) | | 4.00E-02 |
| Schizophrenia | 182559 | 11.59 (8.43-15.94) | 3.0E-51 | 187123 | 9.00 (7.51-10.79) | 1.1E-124 | 1.30 (0.90-1.88) | | 0.16 |
| Anti-anxiety medication | 182559 | 5.17 (4.49-5.96) | 5.5E-114 | 187123 | 5.18 (4.72-5.68) | 0 | 0.99 (0.84-1.18) | | 0.94 |
| Antidepressant medication | 182559 | 5.21 (4.51-6.02) | 1.8E-111 | 187123 | 4.02 (3.71-4.36) | 0 | 1.30 (1.10-1.53) | | 2.0E-03 |
| Antipsychotic medication | 182559 | 1.37 (1.04-1.81) | 2.5E-02 | 187123 | 1.74 (1.38-2.21) | 3.8E-06 | 0.79 (0.55-1.13) | | 0.19 |
| Teenage pregnancy | 182559 | 2.57 (2.13-3.09) | 2.5E-23 | NA | NA | NA | NA | | NA |
| Key stage 3 failure | 132685 | 5.36 (4.56-6.30) | 8.1E-93 | 135082 | 5.01 (4.51-5.56) | 1.1E-201 | 1.05 (0.87-1.27) | | 0.64 |
| Key stage 4 failure | 79490 | 2.72 (2.16-3.43) | 2.2E-17 | 79401 | 3.40 (2.93-3.95) | 5.7E-58 | 0.79 (0.60-1.04) | | 0.089 |
| Key stage 4 low pass | 54909 | 5.09 (3.84-6.74) | 7.8E-30 | 51733 | 4.12 (3.34-5.07) | 3.7E-40 | 1.23 (0.86-1.74) | | 0.25 |
| School absences | 131683 | 3.11 (2.61-3.70) | 7.5E-37 | 132858 | 2.55 (2.30-2.82) | 2.2E-73 | 1.21 (0.99-1.48) | | 0.065 |
| **Variable** | **N** | **Beta(SE)** | **p** | **N** | **Beta(SE)** | **p** | **Beta(SE)** | | **p** |
| GP contacts | 182559 | 0.21 (0.00) | 4.0E-178 | 187123 | 0.10 (0.00) | 0 | 2.12 (0.24) | | 2.6E-19 |
| Outpatient contacts | 182559 | 0.06 (0.00) | 7.3E-109 | 187123 | 0.03 (0.00) | 2.1E-181 | 0.65 (0.07) | | 8.1E-23 |
| Inpatient contacts | 182559 | 0.02 (0.00) | 4.7E-53 | 187123 | 0.01 (0.00) | 2.5E-42 | 0.22 (0.02) | | 7.3E-27 |
| Emergency contacts | 182559 | 0.03 (0.00) | 1.1E-78 | 187123 | 0.01 (0.00) | 0 | 0.21 (0.03) | | 6.5E-14 |
| WIMD* | 178346 | 0.05 (0.00) | 3.25E-17 | 182838 | 0.03 (0.00) | 1.54E-70 | -0.09 (0.06) | | 0.12 |

*At end of study in those aged 18+. WIMD: Welsh Index of Multiple Deprivation (higher = greater deprivation). Outcome variable is coded as 0=non-ADHD & 1=later diagnosed ADHD.
